# Early Measurable Residual Disease Detection after CAR-T is Associated with Poor Outcome Large B-cell Lymphoma Patients

**DOI:** 10.1101/2024.11.27.24318095

**Authors:** Nira Krasnow, Katie Maurer, Catherine Song, Justin Rhoades, Kan Xiong, Andela Crnjac, Timothy Blewett, Lily Gao, Heather Jacene, Reid Merryman, Satyen H. Gohil, Caitlyn Duffy, Liliana I. Guerrero, Jamie Dela Cruz, Mikaela McDonough, Jacquelyn O. Wolff, Robert Redd, Mike Mattie, Brodie Miles, G. Mike Makrigiorgos, Donna S. Neuberg, Scott J. Rodig, Philippe Armand, Caron Jacobson, Viktor A. Adalsteinsson, Catherine J. Wu

## Abstract

Despite responses of chimeric antigen receptor (CAR)-T cells in relapsed/refractory (R/R) large B cell lymphoma (LBCL) patients, over half of patients eventually relapse. Methods to detect early disease persistence are needed to identify patients at high-risk of treatment failure. We recently developed MAESTRO, an ultrasensitive, tumor-informed measurable residual disease (MRD) assay, which can detect parts-per-million (ppm) levels of circulating tumor DNA (ctDNA) using minimal sequencing. We applied MAESTRO to 140 samples from 28 patients (15 durable responders at 12 months, 13 nonresponders) to identify treatment failure following axicabtagene ciloleucel (axi-cel) administered at our institution between 2018 and 2022. Responder and nonresponder patients had similar baseline tumor burden. By 1 week after infusion, responders had marked ctDNA reduction compared to nonresponders, p<0.001. At weeks 2 and 4, responders had ctDNA levels approaching 0 ppm, while nonresponders had persistence of ctDNA, each p<0.001. At day 0, 21% of patients had ctDNA fractions below 0.01%, hence these individuals would not have qualified for ctDNA monitoring with a less sensitive test. Our results confirm feasibility of highly sensitive MRD detection by ctDNA for early identification of patients at high risk of disease progression from axi-cel.

## Introduction

Chimeric antigen receptor T (CAR-T) cells targeting CD19 have transformed the treatment landscape for relapsed/refractory (R/R) large B cell lymphoma (LBCL). Nevertheless, long-term remission is only achieved in ~40% of patients.^1^ Methods for early detection of disease persistence are needed to facilitate identification of patients at high risk of treatment failure and prompt further therapeutic intervention.

Approximately one-third of patients will have up-front response 1 month post-CAR-T by positron emission tomography/computed tomography (PET/CT) imaging with 2-deoxy-2-[18F]fluoro-D-glucose (FDG) but subsequently experience progressive disease.^2^ Highly sensitive ctDNA detection methods may predict longer-term outcome, particularly in this subset of patients with transient responses. Circulating tumor DNA (ctDNA) shows potential for sensitive and early detection of disease persistence or relapse. In patients with lymphoma, ctDNA can be assayed in peripheral blood samples and has already shown promise as a biomarker for detection of patients at high risk of treatment failure after first-line chemo-immunotherapy or after CAR-T. ^3–6^

We recently developed MAESTRO, a highly sensitive, tumor-informed, mutation enrichment sequencing measurable residual disease (MRD) assay, which can detect at or below parts-per-million (ppm) levels of ctDNA using minimal sequencing ^7,8^. Given its high efficiency, bespoke MAESTRO tests can also be pooled and applied to many patients’ samples (MAESTRO-Pool) which confers the advantages of being able to simultaneously assess MRD in patient-matched samples and confirm specificity in patient-unmatched samples.^6,9^ Here, we evaluated whether this method could predict response in patients with R/R LBCL following CD19 CAR-T therapy.

## Methods

All patients provided informed written consent to IRB-approved protocols at the Dana-Farber/Harvard Cancer Center allowing access to clinical data for research purposes and evaluation of clinical samples.

Response was assessed by PET/CT at 1-, 3-, 6-, and 12-months following CAR-T and determined by 2014 Lugano criteria (**Figure 1A**).^10^ Responders (R) were defined as those achieving complete metabolic response (CR) or partial response (PR) continuing through 12 months, while non-responders (NR) were defined as having stable disease (SD) or progressive disease (PD) at any FDG-PET/CT evaluation before 12 months (**Figure 1B,C; Supplemental Figure 1A**). Tumor and normal DNA, isolated from individual tumor tissue and peripheral blood mononuclear cells (PBMC), respectively, were used to create personalized tumor mutation fingerprints. Circulating tumor DNA (ctDNA) was quantified in plasma and/or serum samples from at least 3 timepoints within one month from infusion of CAR-T therapy and, where available, from additional time points to one year.

**Figure 1.**
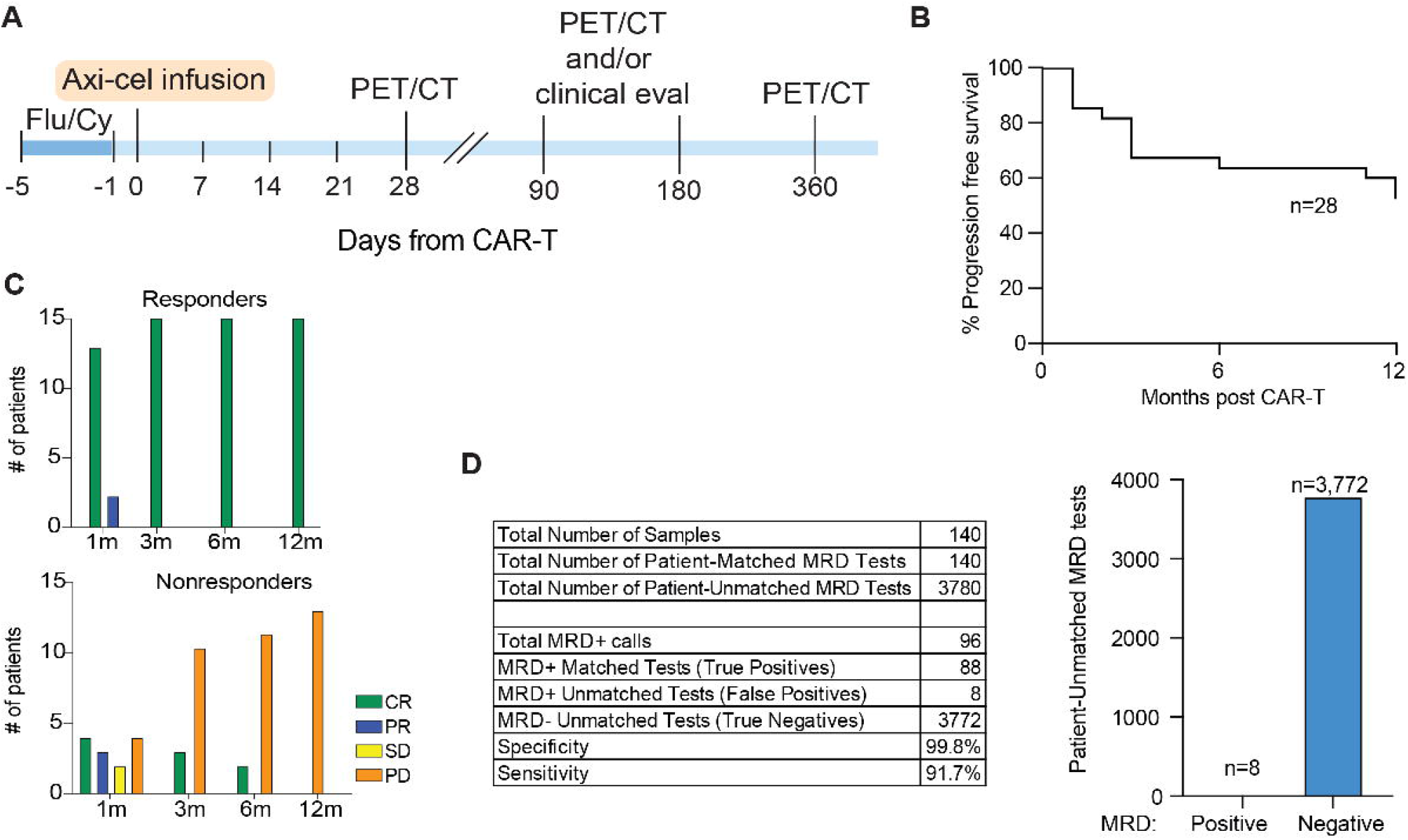
**A)** Schematic of treatment timeline and response assessments. **B)** Kaplan-Meier curve showing progression free survival (PFS) of patients included in the analysis. **C)** Number of patients with complete response (CR. green bars), partial response (PR, blue bars), stable disease (SD, yellow bars), or progressive disease (PD, orange bars) at each response assessment time point by PET/CT scan for patients who are classified as durable responders at 12 months (top) or nonresponders at/before 12 months (bottom). **D)** Table shows MRD detection in patient-matched and -unmatched tests. Bar chart depicts the number of patient-unmatched MRD tests that were positive vs. negative for MRD.

Whole genome sequencing (WGS), sample preparation, and MAESTRO-Pool assays were performed as previously described.^9,11,12^ Briefly, tumor and germline WGS were performed to identify patient-specific tumor somatic single nucleotide variants (SNVs), which were used to design patient-specific MAESTRO probes tracking up to 5000 SNVs per patient. Cell-free DNA (cfDNA) was isolated from patient plasma samples. MAESTRO probes for all patients were pooled together to form a MAESTRO-Pool panel which was applied to all samples from all patients. Tumor fraction (TFx) corresponds to the fraction of cancer-derived versus total cfDNA. After filtering (**Methods**), MRD status, TFx, and limit of detection (LOD95) were calculated as previously described.^9^ LOD95 is the lowest tumor fraction at which ctDNA detection would be predicted to occur with 95% likelihood. This is not the lowest TFx that can be reliably detected. TFx fold changes were computed relative to baseline; when ctDNA was undetectable at a subsequent time point, the LOD95 was substituted for the TFx to reflect a minimum fold change as previously described.^8^

## Results

We evaluated samples from 35 patients with R/R LBCL treated with CAR-T (axicabtagene ciloleucel [axi-cel]) between 2018 and 2022 at our institution (**Table 1**). Responder and nonresponder patients were similar in terms of baseline (pre-bridging) tumor burden (**Supplemental Figure 1B**) and other baseline characteristics, except that NRs had a shorter interval from diagnosis to CAR-T (median 12.8 vs 39.6 months, p=0.008). We applied MAESTRO-Pool to all patient samples, comprising a collection of patient-specific MAESTRO tests targeting 143-5000 SNVs per patient (median: 1895), across 179 cfDNA samples (**Supplemental Figure 1C**). Four patients were excluded due to having undetectable ctDNA at baseline, most likely due to their tests being underpowered with LOD95s exceeding 1000 ppm. Two patients did not have baseline plasma available, and one patient died from non-cancer related causes within one year of CAR-T. (**Supplemental Figure 1D**). Thus, 28 patients were included in the final analysis (15 R, 13 NR). Five patients had PR at first FDG-PET/CT assessment, with 2 converting to CR and 3 to progressive disease before 12 months. Three patients with initial CR subsequently progressed by 1 year. MAESTRO-Pool demonstrated remarkable specificity for individual patient tumor fingerprints; of the 3780 patient-unmatched MRD tests, only 8 were ctDNA-positive, yielding a false positive rate of 0.2% (**Figure 1D**). Notably, one plasma sample for NR01 was responsible for 3 of the 8 false positive calls.

**Table 1:**
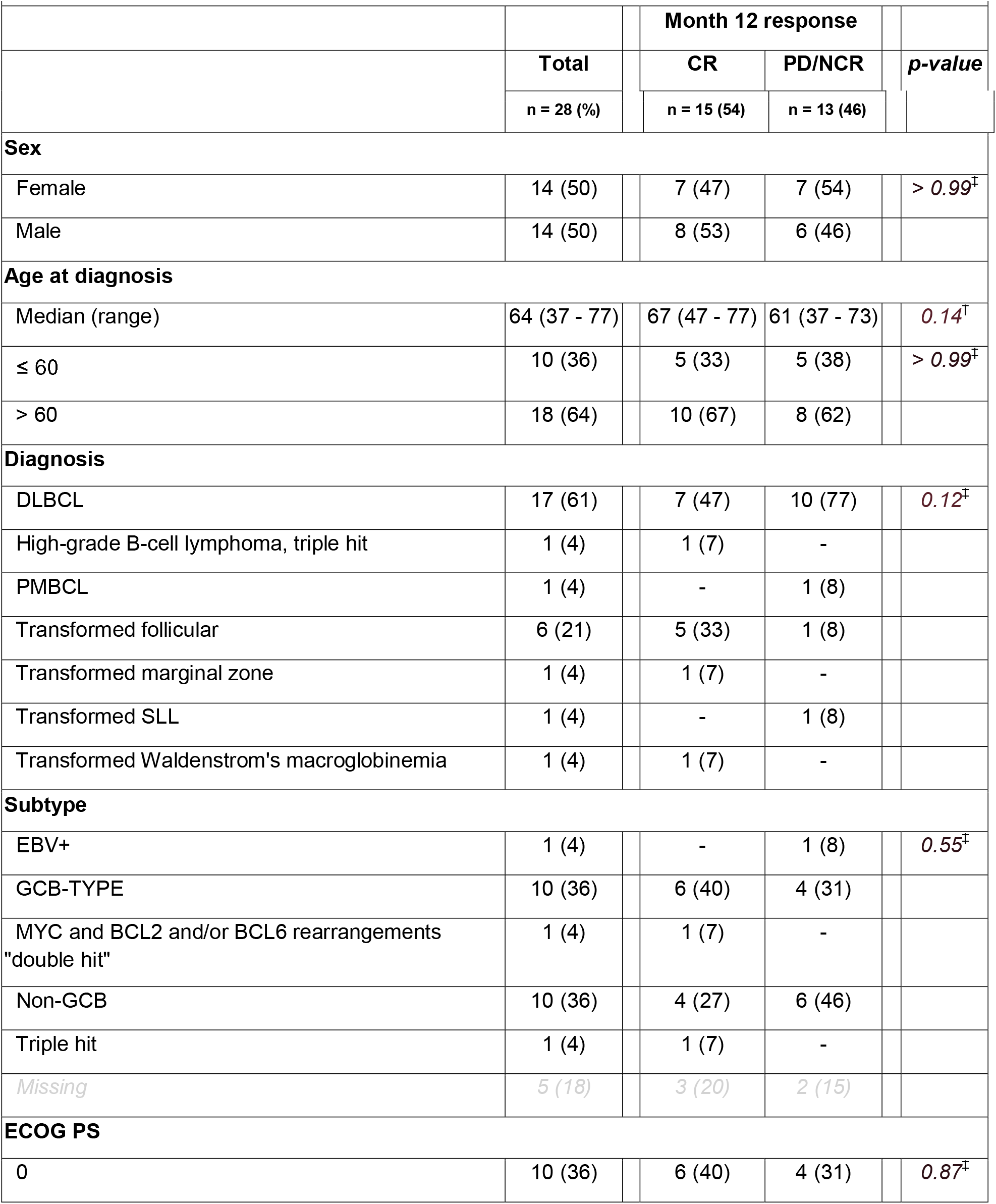
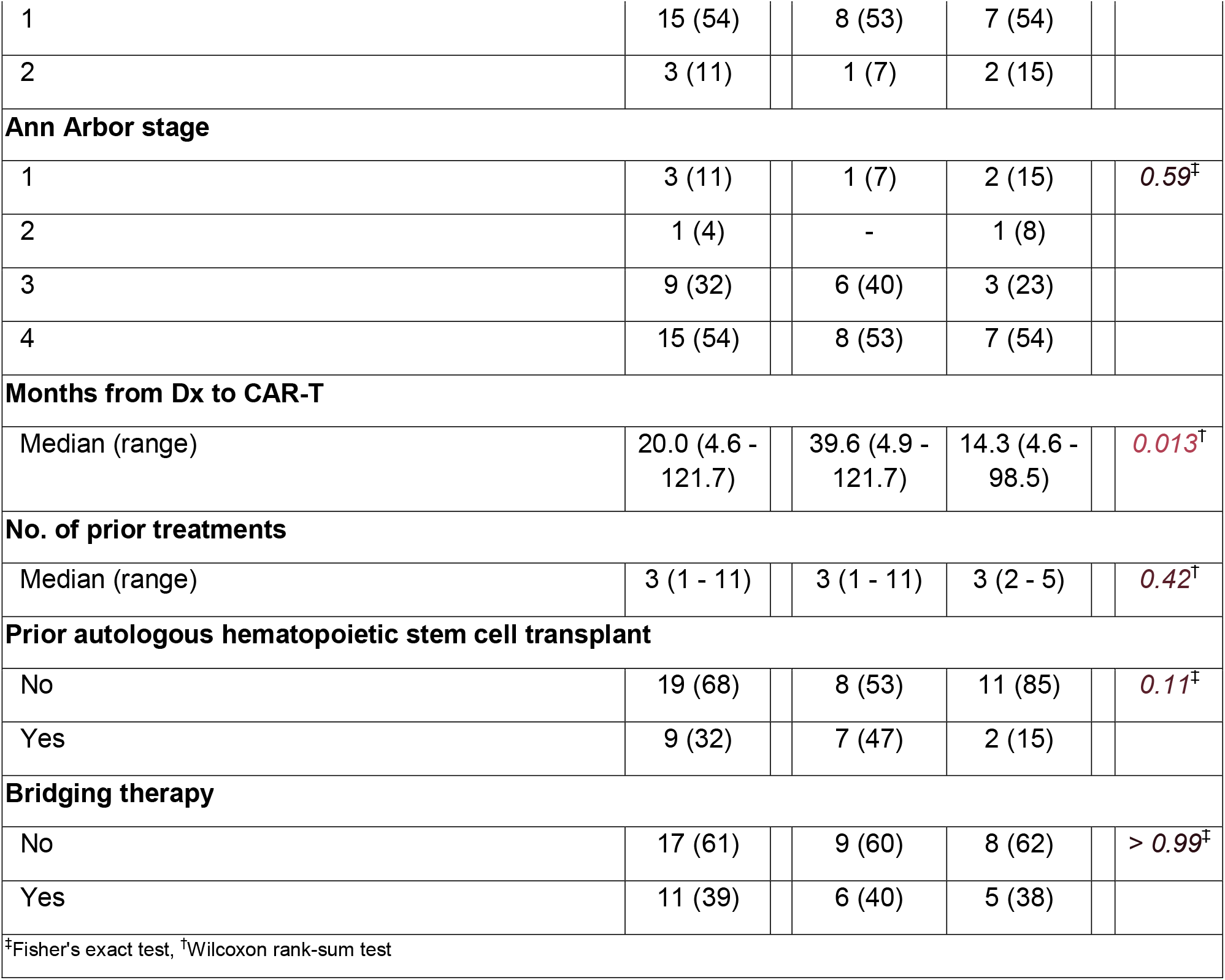
Patient characteristics by month 12 response.

We examined the association of response to CAR-T treatment with ctDNA dynamics from MAESTRO-Pool. All patients had detectable ctDNA at baseline (day 0 i.e. day of CAR-T infusion), with a median TFx of 872.26 ppm (range: 3.92-87361.24 ppm). At baseline, 21% of patients had ctDNA fractions below 100 ppm, and we observed a trend toward an increase in tumor fraction (TFx) in NR compared to R (p=0.07) (**Figure 2A**). At 1 week, all but one patient had detectable ctDNA, with a median TFx of 199.98 ppm (range: 0-104504.29 ppm). At weeks 1 and 2, ctDNA levels were markedly lower in Rs, approaching 0 ppm, compared to NRs (p<0.001 at both timepoints) (**Figure 2A**). By 1 week, Rs demonstrated lower TFx compared to NRs (p<0.001), and the median TFx increased by 2.96-fold for NRs and decreased by 5.90-fold for Rs relative to baseline (p=0.01). By 2 weeks, the TFx decreased by 1.22-fold for NRs and 8.60-fold for Rs relative to baseline (p=0.01). By weeks 3 and 4, the TFx continued to decline, reaching 24.5-fold and 2.58-fold decrease for NRs, respectively; and 76.1-fold and 69.5-fold decrease for Rs, respectively (p=0.24 and 0.06, respectively, **Figure 2B**).

**Figure 2.**
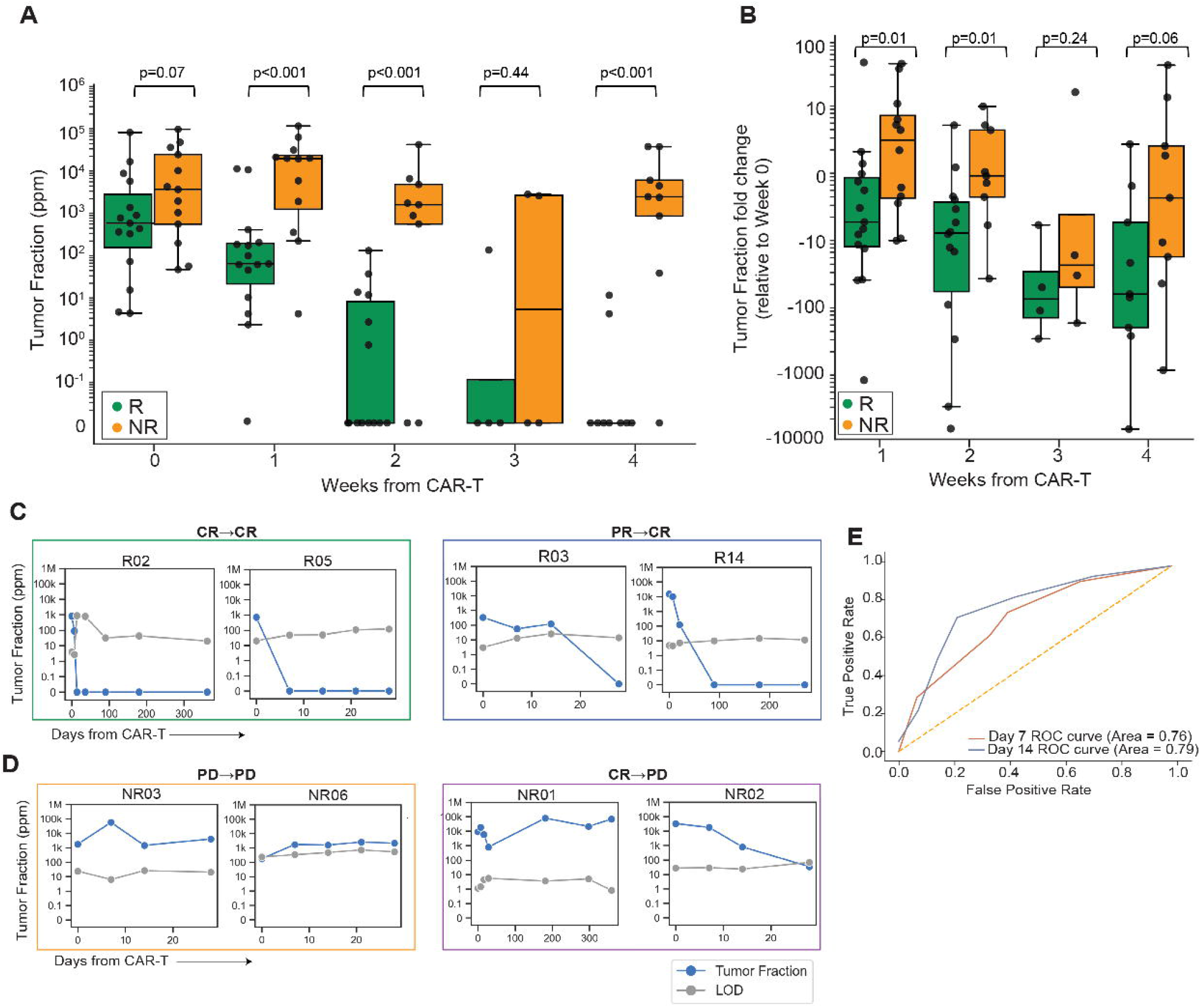
**A)** TFx levels observed during CAR-T treatment categorized by response status. **B)** TFx fold-change observed relative to baseline categorized by response status. For W1 and onward samples with undetectable ctDNA, LOD95 was substituted for TFx to reflect a minimum fold change in TFx. **C)** TFx with corresponding limit of detection (LOD95) for two example patients following CAR-T in two example patients achieving CR at 1 month and maintaining response through 1 year (green box, left) and two patients achieving PR at 1 month and subsequently converting to CR (blue box, right). Of note: LOD95 reflects the lowest tumor fraction at which there is 95% likelihood of detection, as opposed to the lowest detectable TFx. **C)** TFx with corresponding LOD95 for two example patients following CAR-T in two example patients PD at 1 month (orange box, left) and two example patients achieving CR or PR at 1 month and subsequently progressing (purple box, right). For C,D - Blue line indicates TFx and grey line indicates LOD95. ctDNA, circulating tumor DNA; TFx, tumor fraction. **E)** Overall performance of predicting patient response status at Week 1 and Week 2 using TFx fold change from baseline.

Most Rs (n=11 of 15, 73%) had undetectable ctDNA within 4 weeks of treatment (**Supplemental Figure 2**). At a qualitative level, the two patients with initial PR who subsequently converted to CR had ctDNA trajectories similar to patients with upfront CR (**Figure 2C**) whereas patients with initial CR or PR (n=6) who subsequently had PD had overall ctDNA trajectories more akin to patients with up-front PD (**Figure 2D**). This suggests that MRD detection by ctDNA may be a more accurate predictor of durable response at 1 year compared to FDG-PET/CT determination of response although further validation in larger cohorts is needed. Analysis of the receiver operating characteristic curves for TFx and TFx fold change on weeks 1 and 2 revealed ctDNA at week 2 consistently emerging as a stronger predictor of patient response status overall (**Figure 2E, Supplemental Figure 3A**). Indeed, the ROC values were higher at week 2 (AUC=0.97) than at week 1 (AUC=0.87).

To evaluate the impact of bridging therapy on ctDNA dynamics, we examined 11 of the 28 patients (39%) who received bridging therapy between leukapheresis and CAR-T infusion, including 6 Rs and 5 NRs (**Supplemental Table 1**). No significant differences were observed in TFx or TFx fold change between patients who received bridging therapy and those who did not (**Supplemental Figure 3B**,**C**).

For patients with PR at 1 month who later converted to CR, MAESTRO-Pool showed early ctDNA clearance in a similar manner to patients with CR at 1 month (**Supplemental Figure 3D**,**E**). Conversely, the 4 patients with 1 month PR who later progressed did not have ctDNA reduction, similar to patients with up-front PD.

## Discussion

To date, few clinical biomarkers have been identified that predict response to CAR-T therapy.^13,14^ None have yet been successfully applied to standard clinical care to identify patients with LBCL at high risk of relapse to allow for early intervention.

Here, we applied MAESTRO-Pool for tracking ctDNA in patients with relapsed/refractory LBCL as an early indicator of MRD and disease progression. This approach is both highly sensitive and specific, identifying differences in MRD as early as 1 week post-infusion that strongly associate with durable 12-month response to CAR-T. Within one week of CAR-T infusion, eventual responders demonstrated a marked reduction in ctDNA, whereas patients who went on to have progressive disease demonstrate persistence of ctDNA. Our results suggest that ctDNA level at week 2 or fold change from baseline can predict durable response at 1 year. Moreover, ctDNA measurement early after CAR-T infusion may better predict long-term outcome compared with FDG-PET/CT evaluation, particularly for those with PR by FDG-PET/CT at 1 month; however, this observation requires more robust validation in larger cohorts.

Nonresponder patients generally had high TFx that did not decrease with time. However, the high sensitivity of MAESTRO-Pool enabled monitoring of response in more patients and at greater depth than would be detectable using a less sensitive test, suggesting this could be a more viable option for disease monitoring than other less sensitive tests that are currently available. This may also aid in the differentiation between transient and durable responses by 1 week. These findings suggest that ctDNA clearance can predict durable response while ctDNA persistence as a proxy for MRD may be an effective tool for early identification of patients with LBCL at highest risk of disease progression, which could help inform early intervention for these patients in an effort to improve outcomes.

## Data Availability

All data produced in the present study are available upon reasonable request to the authors.

## Acknowledgements

We thank the Ted and Eileen Pasquarello Tissue Bank and Doreen Hearsey for prospective collection and processing of serial serum/plasma samples and the DFCI/HCC cellular therapies support staff for continued care of CAR-T patients. Funding for this study was provided by Kite, a Gilead company, and the Gerstner Family Foundation. K.M. received support from the Lubin Family Foundation. P.A. gratefully acknowledges the support of the Leukemia and Lymphoma Society as well as the Harold and Virginia Lash Foundation. C.J.W is Lavine Family Chair for Preventative Cancer Therapies.

## Author Contributions

K.M., S.H.G., N.K., J.R., K.X., G.M.M., V.A.A., C.J.W., P.A., and C.J. conceived and designed the study. K.M., S.H.G., N.K., R.M., C.D., L.I.G., J.D.C., J.O.W., S.J.R., and M.M., identified, collected, and interpreted patient information. K.M., E.L., K.X., T.B., A.C., L.G., designed and performed experiments. H.J. performed metabolic tumor volume (MTV) quantification. R.R. and D.S.N. designed and performed statistical analysis. K.M., N.K., C.S., J.R. K.X., T.B., J.B., M.M., and B.M. designed, performed, and interpreted data analysis. All authors participated in manuscript writing and review and provided final approval of the manuscript.

