## Supplemental Material for "Early Measurable Residual Disease Detection after CAR-T is Associated with Poor Outcome Large B-cell Lymphoma Patients"

**Supplemental Methods**

**Metabolic tumor volume (MTV) measurement**

For baseline tumor burden estimation measured by metabolic tumor volume (MTV) by PET/CT scan, regions of interest were drawn around tumor using a standardized uptake value (SUV) threshold of 2.5 using MIM software to determine MTV.^11^ For 4 patients (3 R, 1 NR), PET/CT scan prior to CAR-T infusion was not available for analysis.

**MAESTRO site filtering**

Only tumor SNVs that were validated in the tumor tissue and absent in the matched normal blood cells were considered. Additionally, we applied a site-level outlier filter for low-level TFx samples (defined as ≤ 10 ppm). This filter assessed individual fingerprint sites with abnormally high variant allele frequency (VAF); only sites which passed a p-value threshold of ≥ 0.01 were included in downstream analysis.12 TFx fold change was computed for week 1 (W1) to week 4 (W4) samples with respect to the baseline at week 0 (W0).

**Supplemental Table 1**

| **Patient ID** | **Bridging yes/no** | **Type of Bridging** |
| --- | --- | --- |
| R01 | No |  |
| NR01 | No |  |
| R02 | No |  |
| R03 | Yes | XRT, Steroids |
| NR02 | No |  |
| NR03 | Yes | Steroids |
| R04 | No |  |
| NR04 | No |  |
| R05 | Yes | Steroids |
| NR05 | Yes | RGemOx+MTX, ibrutinib |
| R06 | No |  |
| NR06 | Yes | BR and polatuzumab vedotin, XRT |
| R07 | No |  |
| R08 | No |  |
| NR07 | No |  |
| NR08 | No |  |
| NR09 | Yes | R-DHAC |
| R09 | Yes | Rituximab and polatuzumab vedotin |
| R10 | No |  |
| NR10 | No |  |
| R11 | Yes | Rituximab and polatuzumab vedotin, steroids, XRT |
| R12 | No |  |
| NR11 | No |  |
| R13 | Yes | Steroids |
| NR12 | Yes | ibrutinib + Steroids, RGemOx |
| R14 | Yes | Steroids |
| NR13 | No |  |
| R15 | No |  |
| Abbreviations: XRT - radiation therapy; R-GemOx- Gemcitabine-oxaliplatin plus rituximab; MTX - methotrexate; BR - Bendamustine and rituximab; R-DHAC - Dexamethasone, High dose Ara-C (Cytarabine), and Carboplatin; R-pola - Rituximab and polatuzumab vedotin | | |

**Supplemental Figure legends**

**Supplemental Figure 1. Cohort summary and patient outcomes. A)** Swimmer plot of patient responses after CAR-T. **B)** Lymphoma burden at baseline measured by metabolic tumor volume (MTV) from PET/CT scan preceding CAR-T. P value shown by Wilcox. **C)** MAESTRO-Pool matrix of MRD detection for patient-matched MRD tests on the diagonal indicating TP detections and off-diagonal signals indicating patient-unmatched MRD tests with FP detections. Intensity of TFx for MRD-positive tests is visually represented by the color scale. MRD, minimal residual disease; TP, true positive; FP, false positive, TFx, tumor fraction. **D)** Diagram of included and excluded patients in this study.

**Supplemental Figure 2. Longitudinal monitoring of ctDNA TFx and associated LOD.** TFx with corresponding LOD95 following CAR-T for responder (top) and nonresponder (bottom) patients. Blue line indicates TFx and grey line indicates LOD95. Plots in the red box represent patients who have an initial CR or PR and subsequently have PD. ctDNA, circulating tumor DNA; TFx, tumor fraction; LOD, limit of detection. LOD95 represents the lowest TFx at which we are 95% powered to detect ctDNA for a given sample, as opposed to the lowest detectable TFx. In other words, MAESTRO can still detect ctDNA below the LOD95.

**Supplemental Figure 3. ctDNA dynamics correlated with response status**. **A)** Overall performance of predicting patient response status at Week 1 and Week 2 using TFx. **B)** TFx levels and **C)** TFx fold-change observed relative to baseline categorized by receipt of bridging therapy. Statistical significance was evaluated using the Mann-Whitney *U* test. ctDNA, circulating tumor DNA; TFx, tumor fraction; W, week. **D)** TFx levels and **E)** TFx fold-change relative to baseline observed during CAR-T treatment categorized by response status. Response status is further divided into stable (Responder, Non-Responder) and transitory groups (Responder_Non-Responder).

**Supplemental Figure 1**

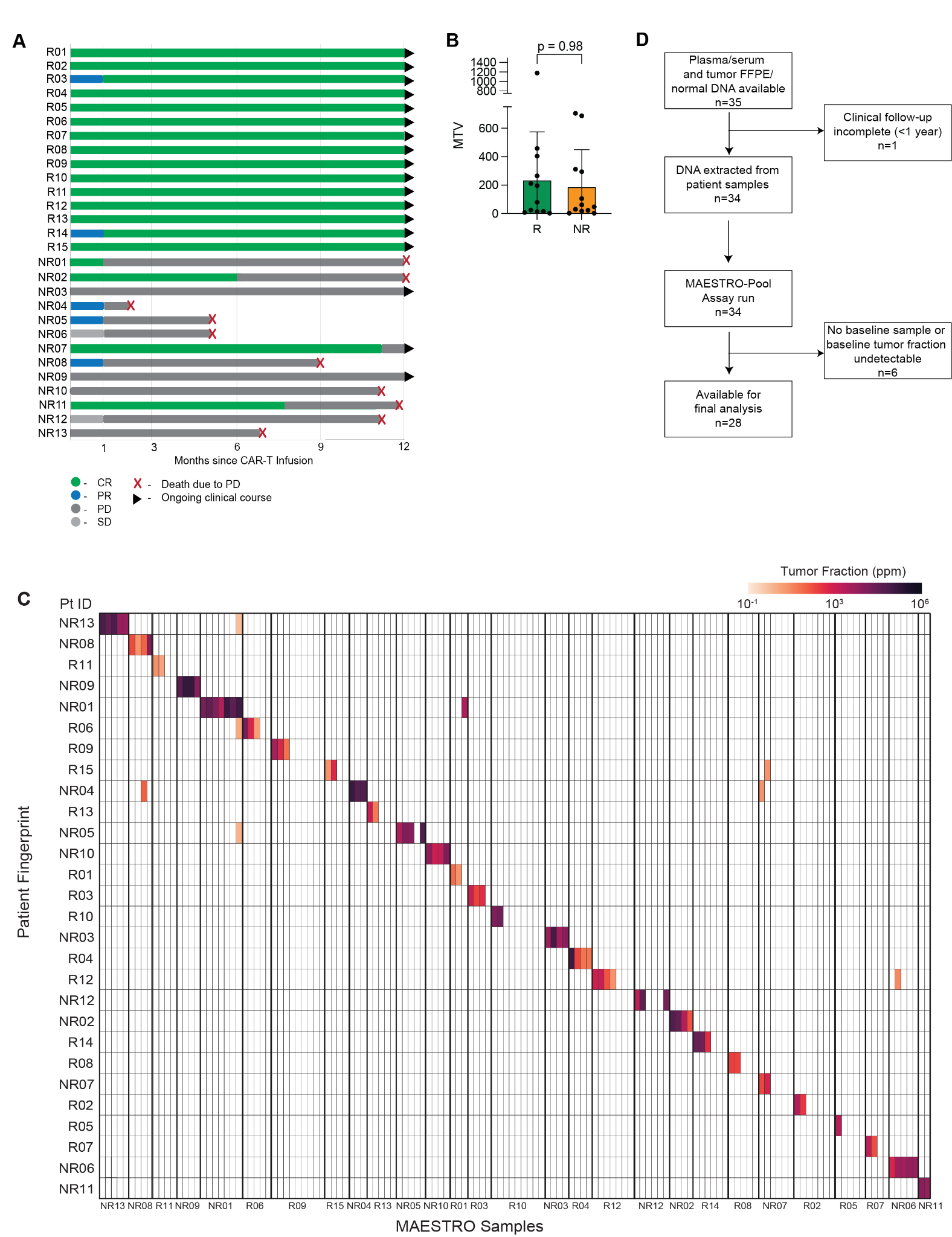

**Supplemental Figure 2**

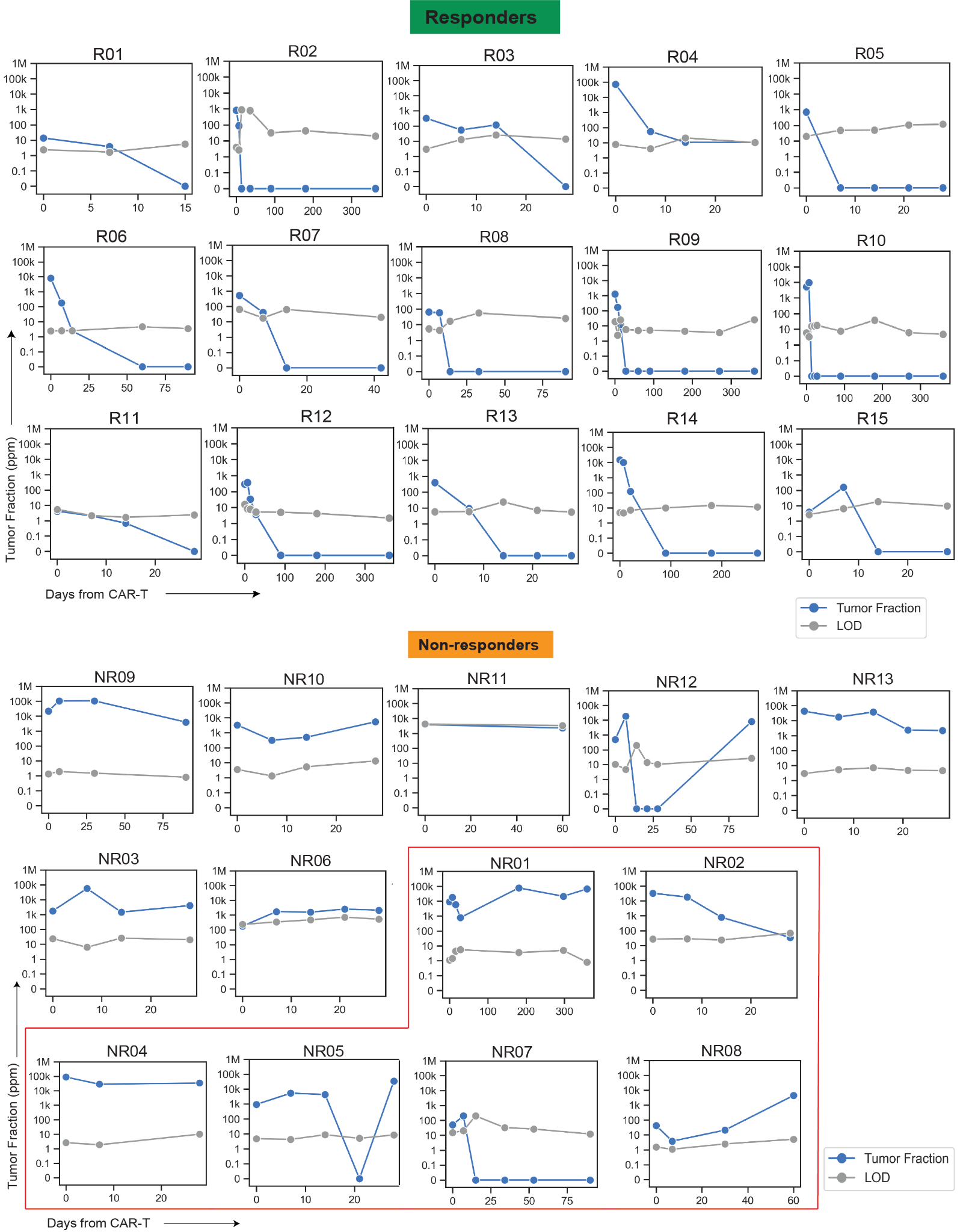

**Supplemental Figure 3**

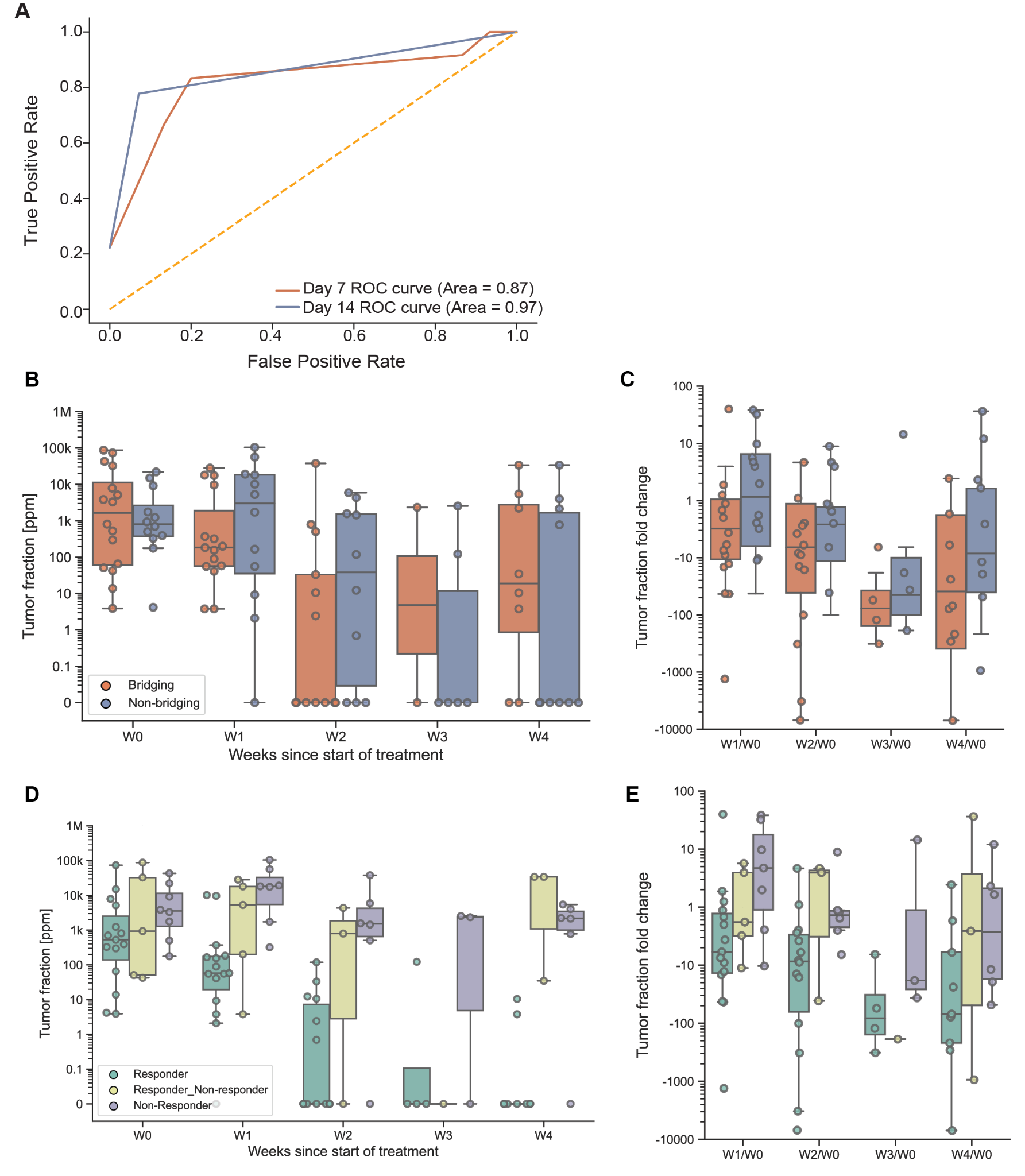
